# Genetic Underpinnings of Regional Adiposity Distribution in African Americans: Assessments from the Jackson Heart Study

**DOI:** 10.1101/2020.10.27.20220517

**Authors:** Mohammad Y. Anwar, Laura M. Raffield, Leslie A. Lange, Adolfo Correa, Kira C. Taylor

## Abstract

**Background:** African ancestry individuals with comparable overall anthropometric measures to Europeans have lower abdominal adiposity. To explore genetic underpinning of different adiposity patterns, we investigated if genetic risk scores for well-studied adiposity phenotypes also predict other adiposity measures in 2420 African American individuals from the Jackson Heart Study.

**Methods:** Polygenic risk scores (PRS) for BMI, WHR adjusted for BMI (WHR_BMIadj_), WC_BMIadj_, and body fat percentage (BF%) were calculated using GWAS significant variants from mostly European ancestry studies. Associations between each PRS and adiposity measures were examined using multivariable linear regression.

**Results:** The BMI-PRS was found to be a positive predictor of BF% (β=0.005 per allele, 95% CI: 0.002, 0.008) and subcutaneous adiposity (β=0.004, CI: 0.002, 0.008). The BF%-PRS was associated with subcutaneous (β=0.022, CI: 0.010, 0.032) but not visceral adiposity; neither BMI nor BF%-PRS were predictors of central obesity measures. Other PRS were not associated with BF%.

**Conclusion:** These analyses suggest: (a) genetically driven increases in BF% strongly associate with subcutaneous but not visceral adiposity; (b) BF% is strongly associated with BMI but not central adiposity associated genetic variants. How these variants may contribute to observed differences in adiposity patterns between African and European ancestry individuals requires further study.

## Introduction

Despite wide adoption of BMI, a measure that correlates well with numerous health risk factors^1^, there are limitations to this metric^2^; notably, it does not differentiate between variation in fat and lean mass. This can lead to imprecise categorization of obesity^3^, and hence misleading inferences for cardiometabolic outcomes^4^, a major drawback given that obesity-health outcome associations are differentially mediated by bodily composition, with adipose tissues more prominently linked with adverse outcomes^5^. Both total adiposity^6^ and regional fat distribution^7^ are significant risk factors for cardiovascular diseases, particularly visceral adiposity^8^.

Since body fat is linearly associated with BMI in sedentary populations^9^, a proportion of genetic variants associated with overall body mass expectedly overlap with loci linked to body fat percentage^10^. However, this BMI-fat mass link is known to exhibit phenotypic variability across ethnicities^11^, which extends to regional distribution of fat tissue as well: in African ancestry (AA) individuals with comparable BMI metrics to those with European ancestry (EA), distribution of visceral adiposity is lower ^12,13^. Paradoxically, the prevalence of cardiometabolic diseases in AA’s, compared to EA’s with similar BMI distributions is higher^14^; beyond environmental factors^15^, genetics may also play a significant role in this and merits a thorough examination.

Despite evidence for AA-specific variants associated with BMI^17^, most SNPs are found to be the same as those first reported in EA individuals^18^. But evidence for generalization of variants associated with other adiposity traits to AAs, including measures of central obesity is limited^19^, and no study has examined replicability of body fat percentage associated variants to AA’s. Since genomic loci associated with body fat percentage are suggested to be more closely aligned with multiple cardiometabolic disease risks than BMI^20^, assessing if genetic variants associated with adiposity patterns, particularly body fat percentage (BF%) in EA, can be extrapolated to AA individuals is an important question, potentially providing suggestive evidence of variants which may contribute to differential distribution of adiposity traits in AA’s, and by extension may contribute to disparities in health risk profile.

The objective of this this study was to assess the utility of known variants associated with anthropometric and adiposity measures discovered in predominantly EA populations to predict BF%, subcutaneous adiposity tissue (SAT), visceral adiposity tissue (VAT), and SAT:VAT ratio (VSR) among AA individuals from Jackson Heart Study (JHS). We also characterized the association of variants with evidence of directional replicability and nominal significance in JHS for their originally reported adiposity measure to other adiposity measures, allowing us to examine the relationships between adiposity measures.

## Methods

### Study Population

The Jackson Heart Study (JHS) (n=5306) is a community based prospective observational cohort study among non-institutionalized African Americans recruited from the three counties that comprise the Jackson metropolitan area in Mississippi. Baseline observations were obtained between 2000-2004, with second and third visits occurring in 2005-2008, and 2009-2012 respectively.

For BMI and WC genome-wide association tests, we used phenotypic observations from visit 1 to maximize sample size (N=3020); for other traits, we used phenotypic measures from visit 2 for the same purpose (N=2554). However, for polygenic risk score regression analyses, we exclusively used phenotypic observations from visit 2. In our analyses, 10 participants were excluded due to pregnancy during the visit 2 examination, and 124 were excluded for missing or biologically implausible recording. The total sample size used was n=2420 individuals.

### Genotyping

It was performed with Affymetrix 6.0 SNP Array (Affymetrix, Santa Clara, Calif). Outliers based on principal components, sample swaps, duplicates, and one of each pair of monozygotic twins were excluded. Variants with a minor allele frequency ≥ 1%, a call rate ≥ 90%, and a Hardy Weinberg equilibrium (HWE) p-value >10^−6^ (n=832,508 variants) were used for imputation to 1000 Genomes Project population SNP reference panel (Phase 3, Version 5), using Minimac3 on the Michigan Imputation Server. Only SNPs with an imputation r^2^>0.9 were selected for polygenic risk score analyses.

### Polygenic Risk Scores

Use of polygenic risk scores (PRS) to predict complex traits^21^, closely related phenotypes^22^, and the same phenotype across different populations^23^ has been validated. Both weighted (i.e. adjusting for variant’s effect size on phenotype) and unweighted (frequency counting) methods have been employed for estimation of PRS^24^. Although the weighted method can lead to reduced mean square error for prediction in some cases^25^, the main applications for polygenic scores, namely association testing and prediction, do not appear to differ substantially between two methods. In addition, an unweighted score is more robust against error in estimating the effect sizes arising from limited samples, “winner’s curse bias”^26^, and confounding by demographic structure^27^. Therefore, we used an unweighted PRS approach.

At each locus (SNP), participants were assigned a dosage value between 0-2 inclusively based on the estimated number and frequency of phenotype increasing alleles under an additive genetic effect model, and the PRS value for each individual reflects the summation of risk alleles across all selected loci.

### SNP Risk Sets

To construct SNP sets used for PRS calculation, we utilized both the European Bioinformatic Institute repository (*ebi*.*ac*.*uk/gwas*), and PubMed for extraction and validation of SNPs linked to anthropometric measures including body mass index (BMI), waist to hip ratio adjusted for BMI (WHR_BMIadj_), waist circumference adjusted for BMI (WC_BMIadj_), and body fat percentage (BF%). Most variants originate from EA studies, some from large multi-ethnic research. PRS derived from only AA-specific variants for target phenotypes were judged to be underpowered due to low numbers available, as well as PRS calculated from SNPs associated with VAT, SAT and VSR.

We conducted linkage disequilibrium (LD) analysis using the LDlink (*ldlink*.*nci*.*nih*.*gov*). If a pair or a group of SNPs were in LD with one another (R^2^≥0.2), we prioritized sentinel variants from larger and more recent studies and selected a single SNP to ensure independence of variants and avoid double counting the same functional locus (site). This set of LD-pruned variants was then used to calculcate PRS.

### Anthropometry Measures

WC was measured at the umbilicus level using a non-elastic tape measurer and rounded to the nearest centimeter; hip circumference (HC) was measured at the level of the widest circumference over the greater trochanter. WHR was obtained by dividing WC over HC.

### Adiposity Measures

A variety of different techniques for assessment of body composition exist^28^. For overall BF%, bioimpedance is a widely adopted method^29^. Under this technique, BF% is calculated based on the measured resistance of the adipose tissue as the person lays supine with electrodes placed on the arm and/or leg; bare foot-to-foot bioimpedance was conducted using the Tanita Body Fat Monitor (Tanita Corp, Tokyo). BF% was estimated using a programmed algorithm that incorporates bioimpedance readings with a height, weight, age and sex-specific equation and additional adjustment for physical activity levels.

To estimate visceral and subcutaneous adipose volumes, the study employed computed tomography (CT) technique at visit 2, where the heart and lower abdomen regions were scanned with 16-channel mutidetector CT machine (Lightspeed 16 Pro, GE Healthcare, Milwaukee, WI). Abdominal imaging slices covering the lower abdomen from L3 to S1 were used to quantify both VAT and SAT^30^, such that 24 adjacent 2-mm thick slices centered on the lumbar disk space at L4 to L5 were used for quantification of both types of adiposity; 12 images before the center of L4 to L5 disk space and 12 images after that space^30^.

### Statistical Analysis

To facilitate comparison across different phenotypes, we performed inverse-normal transformations prior to analysis. Genome-wide association analyses were completed for target traits to assess if known loci for anthropometric and adiposity traits discovered in EA populations are directionally (i.e. allele-effect direction) and statistically significantly replicable in the JHS cohort variants with genome-wide significance level (Supplementary Table 1). We used EPACTS 3.2.6^31^ to perform GWAS analyses, adjusting for age, sex, and a genetic relationship matrix using the EMMAX test; additionally, BMI was incorporated as covariate in WHR and WC analyses.

**Table 1.** Baseline characteristics of the study participants.

| **Variable** | **Total (N=2420)** |
| --- | --- |
| **Male**, N. (%), Unit | 892(36.9%), N |
| **AGE**, Mean (SD), Unit | 60.0(12.4), Year |
| **BMI**, Mean (SD), Unit | 32.2(7.2), Kg/m<sup>2</sup> |
| **BMI Categories** |  |
| Underweight, N. (%) | 26(1.1%) |
| Normal Weight, N. (%) | 244(10.1%) |
| Overweight, N. (%) | 762(31.5%) |
| Obese, N. (%) | 1312(54.2%) |
| Missing, N. (%) | 76(3.1%) |
| **Hip Circumference**, Mean (SD), Unit | 114.7(14.9), cm |
| **Waist Circumference**, Mean (SD), Unit | 102.9(16.2), cm |
| **Waist/Hip**, Mean (SD), Unit | 89.8(8.8), ratio×100 |
| **Adiposity Composition** |  |
| **Fat Mass**, Mean (SD), Unit | 76.3(33.5), kg |
| **VAT**, Mean (SD), Unit | 839.4(383.1), cm<sup>3</sup> |
| **SAT**, Mean (SD), Unit | 2335.8(1014.7), cm<sup>3</sup> |
| **Body Fat Mass**, % | 38.2(9.9), Fat/Total Body Mass × 100 |

**VAT:** Visceral Adipose Tissue, **SAT:** Subcutaneous Adipose Tissue

We calculated PRS under three separate but complementary scenarios using the following configurations: (a) set of all known loci (LD-pruned) reported at genome-wide significance (<5×10^−8^) in multi-ethnic or European studies regardless of replicability in JHS; (b) the subset of risk loci with evidence of directional replication in the JHS-GWAS results; and (c) a more restricted subset of risk loci with evidence for both directional replication as well as nominal statistical significance (p<5×10^−2^) in JHS results (Supplementary Table 2). PRS for BMI, WHR_BMIadj_,WC_BMIadj_, and BF% were first tested against their respective phenotypes in JHS to ensure the validity of constructed predictors (Supplementary Table 3).

**Table 2.**
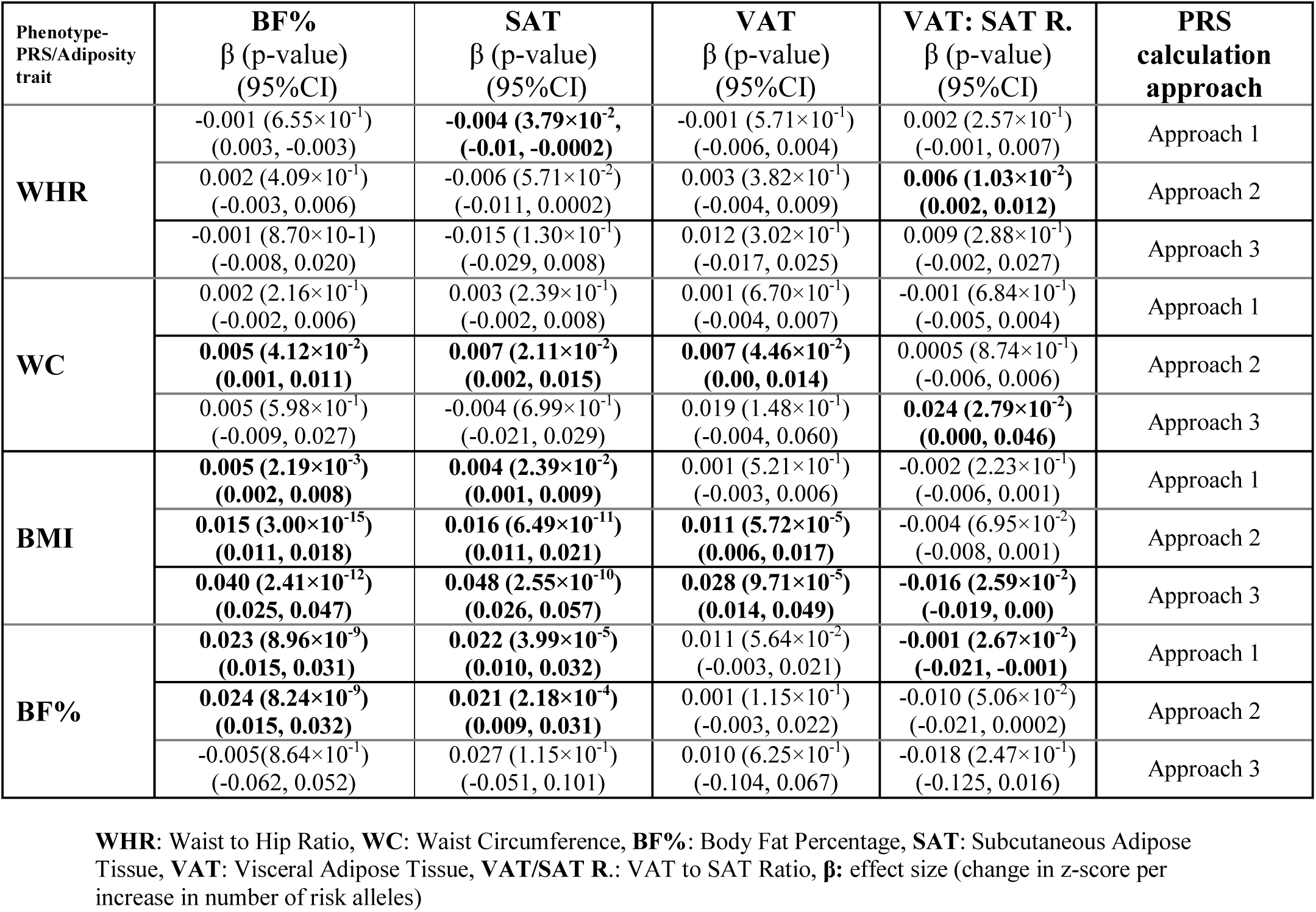
Associations between phenotype-PRS (columns), and measures of adiposity (rows). Betas are reported for standardized inverse normalized values, followed by their respective p-values. Nominally statistically significant results (p<**5**.**00×10**^**-2**^) are in bold font.

Phenotype-specific PRS obtained under various approaches were then tested for cross-sectional associations with phenotypic measures. Both multivariable linear regression and mixed model^32^ were employed to investigate the associations between PRS and adiposity measures, with age, sex, and the top 10 ancestry principal components as covariates; additionally, family ID was utilized as random component in mixed models. Both offered similar results; linear results were more robust and hence chosen for simplicity.

Some loci associated with obesity traits and/or fat distributions^33^ are known to be sex-specific. Although we performed gender-stratified analysis, given the sex imbalance in this sample (36.9% males), results for the females largely mirrored the combined findings, while the male-only analysis lacked precision; therefore we chose to report the sex-combined analysis.

Finally, to characterize the association between EA-established variants for anthropometric traits with evidence of transferrability to AA (i.e. statistically significant in JHS-GWAS), and type of adiposity (BF%, SAT, VAT, VSR), we constructed heatmap plots to investigate if genetic variations underpinning obesity traits are closely correlated with overall body fat change or aligned to specific adiposity patterns.

Statistical results, plots, and heatmaps plots were obtained using RStudio (V 1.1.463), gplots^34^, and “R Color Brewer” packages^35^, respectively.

## Results

### Population characteristics and adiposity measures

Table 1 provides a descriptive distribution of demographic, anthropometric and adiposity traits in the JHS population. Using the standard BMI cutpoints of ≥25 and ≥30 for overweight and obesity respectively, the majority of participants were either overweight (31.5%) or obese (54.2%). The mean WC and BF% also indicate a high prevalence of excess adiposity.

Spearman correlation of anthropometric and adiposity measures (Supplementary Table 4) illustrated a high degree of correlation of BMI with WC (r= 0.82), but much weaker correlation with WHR (r=0.16). BMI is also highly correlated with SAT (r=0.83) and BF% (r=0.70), but not as strongly with VAT (r=0.49).

### Association with Polygenicc Risk Scores

For BF% phenotype, the BF%-PRS was found to be a significant positive predictor under all scenarios (β=0.023 (p=8.96×10^−9^), and 0.024 (p=8.24×10^−9^)) per increase of 1 risk allele under approaches 1 and 2, respectively. The BMI-PRS was found to be positively predictive of BF% in all scenarios (β=0.005 (p=2.19×10^−3^), 0.015 (p=3.00×10^−15^), 0.040 (p=2.41×10^−12^)). However, the PRS for WC_BMIadj_ was only a borderline signficant predictor of BF% under approach 2 (β=0.005(p=4.12×10^−2^)), and WHR_BMIadj_-PRS were not associated with BF% under any approach.

For the SAT phenotype, the BMI-PRS were positive predictors under all three approaches (β=0.004 (p=2.39×10^−2^), β=0.016 (p=6.49×10^−11^), and β=0.048 (p=2.55×10^−10^)); the BF%-PRS were also associated with SAT under the first two approaches (β=0.022 (p=3.99×10^−5^), and β=0.021 (p=2.18×10^−4^)). For the other two phenotypes, the association was only significant under approach 2 for WC_BMIadj_, and approach 1 for WHR_BMIadj_, indicative of poor predictive applicability of known EA central obesity associated SNPs for SAT among AA’s.

For the VAT and VSR phenotypes, the patterns of association were even more inconsistent. While BMI-PRS were predictors of VAT under approach 2 and 3, but only negatively associated with VSR under approach 3. For WC_BMIadj_-PRS, the association with VAT barely reached nominal significance with approach 2, and with VSR under approach 3. For WHR_BMIadj_-PRS, the association with VSR was only nominally significant under approach 2, but never with VAT. BF%-PRS were not predictive of either VAT or VSR under any configuration.

### Association with Individual SNPs

Since PRS were summary statistics of risk alleles and do not illustrate the scale and association of each risk allele may vary when compared to other variants, we performed separate multivariate regression tests with individual variants. We used 20, 10, 15, and 4 risk allele SNPs which were nominally significant in BMI, WC_BMIadj_, WHR_BMIadj_, and BF% GWAS results in JHS, to characterize individual variants’ association with BF%, SAT, VAT, and VSR ratio, respectively; each SNP represents an independent (funcitonal) locus within the genome.

On the heatmap (Figure 1.), the primary clustering of adiposity pattern variables on the x-axis separate different types of fat. The primary clustering of the SNPs on the y-axis separate group of SNPs that are associated with increase in BF%, BMI, WC_BMIadj_, and WHR_BMIadj_ (Figure 1A through 1D, respectively). A clear majority of risk alleles associated with increased BMI and BF% are positively associated with SAT and overall BF%. Among WC_BMIadj_-increasing alleles, the majority are positively associated with VAT and VSR. However, WHR_BMIadj_-increasing alleles appear to be poor predictors of adiposity type in this cohort given no discernible pattern (Figure 1D).

**Figure 1.**
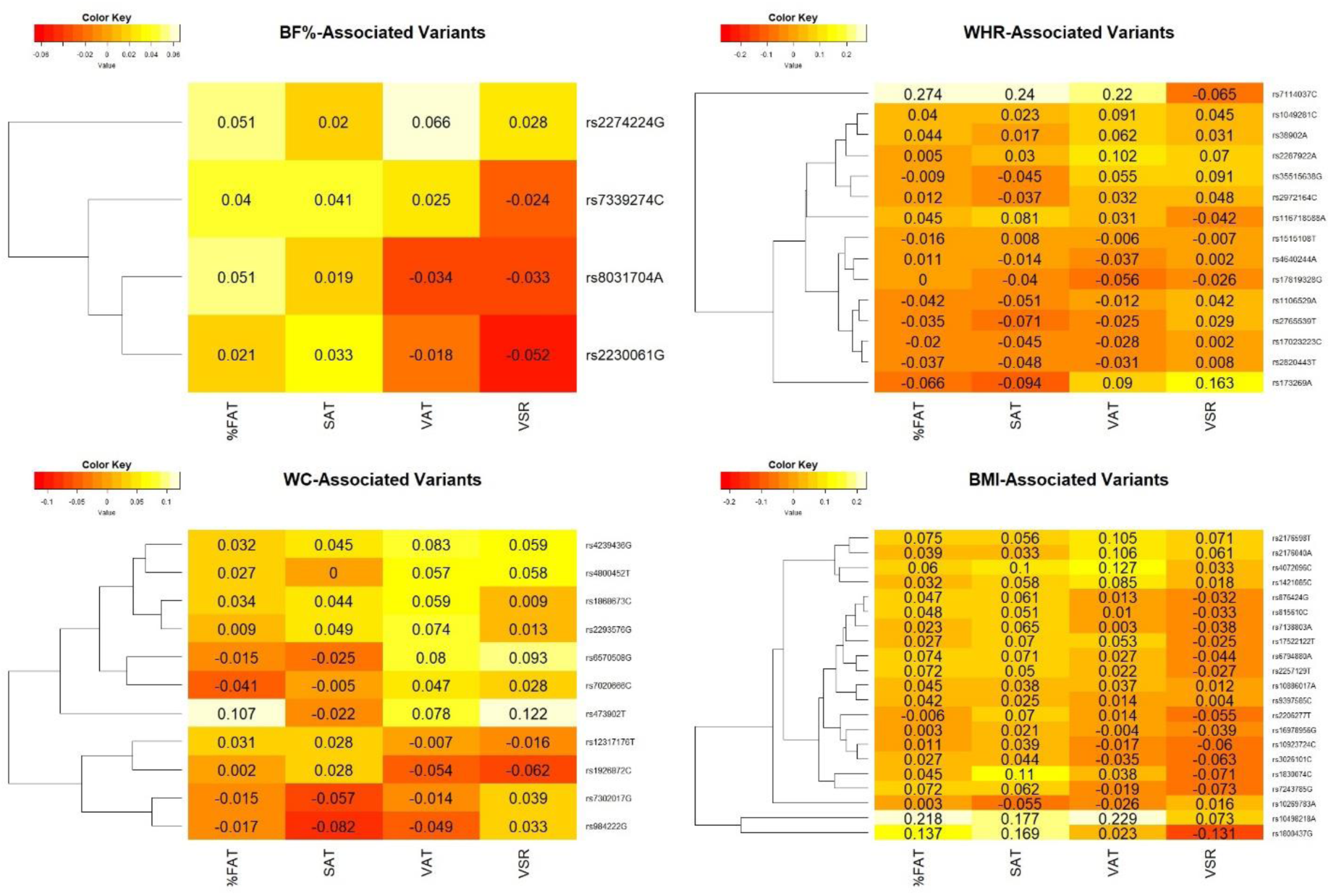
Heatmap plot of association between top phenotype-linked SNPs (represented by phenotype increasing alleles), and adiposity traits. These SNPs were used for PRS calculation under approach 3 and represent polymorphisms with evidence of directionally consistent and statistically significant associations with their respective traits in JHS-genome wide assessment. Abbreviations: **WHR**: Waist to Hip Ratio, **WC**: Waist Circumference, **BF%**: Body Fat Percentage, **SAT**: Subcutaneous Adipose Tissue, **VAT**: Visceral Adipose Tissue, **VSR**: VAT to SAT Ratio.

## Discussion

In this study we characterized the associations between genetic risk scores for BMI, WC_BMIadj_, WHR_BMIadj_, and BF% with body fat composition phenotypes, including overall BF%, VAT, SAT, and VSR among AA individuals in the JHS cohort.

Both BF%-PRS and BMI-PRS were significant and positive predictors of BF% in AA’s; this illustrates that similar to most BMI-linked SNPs^18^, most known BF% associated loci, predominantly observed in EA individuals^20^,, are likely transferrable to AA’s. Furthermore, BMI-associated loci have cross-phenotypic effects on body fat^20^; the significant association between BMI-PRS and BF% measures in our analysis may also be indicative of transferrability of similar biological mechanisms from EA’s to AA’s, though the causal SNPs at most of these loci are unknown.

However, WC_BMIadj_ and WHR_BMIadj_-PRS were not consistently associated with BF% in adjusted analyses which suggests that BF% does not assess adiposity distribution and therefore is unlikely to associate with SNPs for BMI-adjusted central obesity measures. Whether such a poor association between central obesity associated SNPs and BF% is specific to AA’s or similarly generalizable to other groups as well require further studies. Examination of individual SNPs that exhibited nominally significant associations to BF%, BMI, WC_BMIadj_, and WHR_BMIadj_ in this cohort (Figure 1) shows that larger proportions of BMI and BF% associated SNPs cluster with BF% and subcutaneous adiposity measures. Though WC_BMIadj_- associated SNPs cluster with visceral and VSR measures, the pattern is less evident; WHR_BMIadj_- associated SNPs, in contrast, do not appreciably cluster with any of the adiposity traits.

Significant associations between SAT measures and BMI and BF%-PRS may suggest that genetic and/or molecular mechanisms underpinning the variation in subcutaneous adiposity in AA’s are comparable to EA’s; however, similarly it is early to suggest that SAT associated loci in EA populations^36^ may likewise underpin subcutaneous fat configuration in AA’s before well-powered GWAS for SAT in AA’s would become available.

Difference in patterns of visceral adiposity in AA’s^37^, with significantly lower levels of VAT in BMI-adjusted analysis is already noted^13^ that suggests a favorable visceral adiposity profiles compared to EA’s^38^. Due to limited availability of GWAS on VAT or VSR phenotypes^36^, with none in a well-powered AA cohort, it is therefore premature to suggest whether observed differences are primarily genetically driven. AA individuals with comparable visceral fat measures to EA’s have been shown to have higher levels of inflammation biomarkers^39^, which may strengthen the view that genetic mechanisms underpinning visceral fat variation in AA’s could be dissimilar to EA’s. Similar dilution has been observed for closely related phenotypes in some prior multi-ethnic analyses^40^. Larger studies in AA populations are needed to assess these different propositions.

It is the first study to examine the relationship between adiposity PRS, including BF%, WC_BMIadj_ and WHR_BMIadj_, and adiposity traits in AA’s. This is one of relatively few available assessments examining if the genetic architecture underpinning adiposity traits (other than BMI) in AA individuals can be inferred from EA studies. In addition to increased SNP numbers used for PRS calculations that improved precision, utilization of several PRS with different risk set configurations made it possible to examine associations under comparative scenarios, minimizing potential sources of bias in estimates.

Results of this assessment pertain only to adults. Many genetic variants associated with obesity traits are age-dependent^41^. For example, in one prior study, there was no associaiton between BMI-PRS and BF% in <5 years olds^42^; thus, extrapolation of results from this study to younger age categories in AA individuals may be unwarranted. This limitation is likely to be extended to WC_BMIadj_ and WHR_BMIadj_ associated SNPs since they also exhibit interaction with age^43^.

We could not account for differential associations of dimorphic or sex-specific vairants because of smaller male numbers. Bioimpedance have limitations^44^, where the method slightly underestimates BF% in highly obese males ^45^. However, we do not expect this limitation to substantially bias our results since only 6% of males had BMI measures >40 in this dataset. Finally, the cross-sectional nature of the assessment restricts causal inferences.

In conclusion, these analyses illustrate that BMI and BF% associated loci initially explored in predominantly EA populations are generally transferrable to AAs. Our results suggest that total gain in fat mass in AA’s, at least for gains in fat mass mediated by genetic factors, may be mostly through subcutaneous rather than visceral adiposity, but a comparable assessment in other populations, including Europeans, would be required to make firm conclusions. Absence of association between anthropometric PRS, particularly WC_BMIadj_ and WHR_BMIadj_, and adiposity traits may imply that the latter phenotypic measure are likely driven by genetic variants which influence overall adiposity versus central obesity.

## Data Availability

The data that support the findings of this study are available from the corresponding author (MYA), as well as Jackson Heart Study and will be provided upon reasonable request. Codes used for statistical analysis are available from the corresponding author (MYA) and will be provided upon request.

https://www.ncbi.nlm.nih.gov/projects/gap/cgi-bin/study.cgi?study_id=phs000286.v6.p2

## Acknowledgements

The Jackson Heart Study (JHS) is supported and conducted in collaboration with Jackson State University (HHSN268201800013I), Tougaloo College (HHSN268201800014I), the Mississippi State Department of Health (HHSN268201800015I) and the University of Mississippi Medical Center (HHSN268201800010I, HHSN268201800011I and HHSN268201800012I) contracts from the National Heart, Lung, and Blood Institute (NHLBI) and the National Institute on Minority Health and Health Disparities (NIMHD). The authors also wish to thank the staffs and participants of the JHS.

## Disclaimer

The views expressed in this manuscript are those of the authors and do not necessarily represent the views of the National Heart, Lung, and Blood Institute; the National Institutes of Health; or the U.S. Department of Health and Human Services.

