## Supplementary Table 1 for "Genetic Underpinnings of Regional Adiposity Distribution in African Americans: Assessments from the Jackson Heart Study"

**Supplementary Table 1.** GWAS significant results:

| Chromosome | Position | RS# | Nearest Gene | Increasing Allele | MAF | PVALUE | Phenotype |
| --- | --- | --- | --- | --- | --- | --- | --- |
| 3 | 111452110 | rs114130295* | PLCXD2 | G | 0.05612 | 1.44E-08 | BMI |
| 3 | 111454104 | rs4309695* | PLCXD2 | C | 0.05621 | 2.11E-08 | BMI |
| 3 | 111488488 | rs6438011* | PLCXD2 | A | 0.05136 | 1.31E-08 | BMI |
| 10 | 88939181 | rs532236098* | FAM35A | T | 0.00044 | 3.63E-08 | WC |

*not reported in literature for corresponding phenotypes. It is likely these variants are false positives, as they were not reported in much larger meta-analyses.
