## Supplementary Table 2 for "Genetic Underpinnings of Regional Adiposity Distribution in African Americans: Assessments from the Jackson Heart Study"

**Supplementary Table 2.** SNPs configurations used for calculation of polygenic risk scores under approaches 1 to 3.

| Phenotype | All SNPs | Replicated SNPs (for Approach 2) * | Nominally significant (for Approach 3) * |
| --- | --- | --- | --- |
| BMI | 371 | 222 | 21 |
| WHR | 265 | 125 | 15 |
| WC | 189 | 121 | 11 |
| % Fat Mass | 53 | 46 | 4 |

*numbers in the columns 3 and 4 (from the left) represent subset of numbers from preceding columns. Abbreviations: **WHR**: Waist to Hip Ratio, **WC**: Waist Circumference, **BF%**: Body Fat Percentage, **SAT**: Subcutaneous Adipose Tissue
