## Supplementary Table 3 for "Genetic Underpinnings of Regional Adiposity Distribution in African Americans: Assessments from the Jackson Heart Study"

**Supplementary Table 3.** Polygenic Risk Score Validation (phenotypes other than percentage body fat (%BF)):

| Phenotype PRS vs. phenotype measures | Approach 1, β(p-value) * | Approach 2, β(P-value) | Approach 3, β(P-value) |
| --- | --- | --- | --- |
| BMI | **0.008(2.79×10^-5^)** | **0.023(2.00×10^-16^)** | **0.059(2.00×10^-16^)** |
| WHR | **0.005(8.49 ×10^-3^)** | **0.018(2.46×10^-13^)** | **0.063(1.08×10^-13^)** |
| WC | **0.005(5.93×10^-2^)** | **0.011(8.56×10^-4^)** | **0.018(9.96×10^-2^)** |

*reported coefficients and p-values represent estimated change in inverse normalized z-score(s) in Jackson Heart Study phenotype measures per 1 unit increase in polygenic risk score. Estimates are obtained linear regressions models. (Sample Size: N=2420). Abbreviations: **WHR**: Waist to Hip Ratio, **WC**: Waist Circumference, **BF%**: Body Fat Percentage, **SAT**: Subcutaneous Adipose Tissue, **β:** effect size (change in z-score per increase in number of risk alleles)
