## Supplementary Table 4 for "Genetic Underpinnings of Regional Adiposity Distribution in African Americans: Assessments from the Jackson Heart Study"

**Supplementary Table 4.** Matrix of correlation between observed phenotypic measures for Jackson Heart Study. Spearman correlation coefficients were calculated.

| **Spearman correlation coefficient, r (p-value)** | BMI | Waist Circumference | Waist to Hip Ratio | Body Fat % | Subcutaneous Fat Tissue | Visceral Fat Tissue | Visceral: Subcutaneous Fat Ratio |
| --- | --- | --- | --- | --- | --- | --- | --- |
| BMI | NA |  |  |  |  |  |  |
| Waist Circumference | 0.82  (2.20×10^-16^) | NA |  |  |  |  |  |
| Waist to Hip Ratio | 0.16  (2.66×10^-15^) | 0.57  (2.20×10^-16^) | NA |  |  |  |  |
| Body Fat % | 0.70  (2.20×10^-16^) | 0.49  (2.20×10^-16^) | -0.13  (1.72×10^-10^) | NA |  |  |  |
| Subcutaneous Fat Tissue | 0.83  (2.20×10^-16^) | 0.65  (2.20×10^-16^) | -0.04  (9.08×10^-2^) | 0.81  (2.20×10^-16^) | NA |  |  |
| Visceral Fat Tissue | 0.49  (2.20×10^-16^) | 0.63  (2.20×10^-16^) | 0.49  (2.20×10^-16^) | 0.29  (2.20×10^-16^) | 0.33  (2.20×10^-16^) | NA |  |
| Visceral: Subcutaneous Fat Ratio | -0.28  (2.20×10^-16^) | -0.02  (4.96×10^-1^) | 0.45  (2.20×10^-16^) | -0.45  (2.20×10^-16^) | -0.55  (2.20×10^-16^) | 0.54  (2.20×10^-16^) | NA |
